# Classifying Radiology Abstracts with Deep Learning

**DOI:** 10.1101/2021.08.07.21261750

**Authors:** Hongyu Chen, George Shih

## Abstract

**Background:** The Radiological Society of North America (RSNA) receives more than 8000 abstracts yearly for scientific presentations, scientific posters, and scientific papers. Each abstract is assigned manually one of 16 top-level categories (e.g. “Breast Imaging”) for workflow purposes. Additionally, each abstract receives a grade from 1-10 based on a variety of subjective factors such as style and perceived writing quality. Using machine learning to automate, at least partially, the categorization of abstract submissions can result in saving many hours of manual labor.

**Methods:** A total of 45527 RSNA abstract submissions from 2014 through 2019 were ingested, tokenized, and pre-processed with a standard natural language programming protocol. A bag-of-words (BOW) model was used as a baseline to evaluate two more sophisticated models, convolutional neural networks and recurrent neural networks, and also evaluate an ensemble model featuring all three neural networks.

**Results:** ensemble model was able to achieve 73% testing accuracy for classifying the 16 top-level categories, outperforming all other models. The top model for classifying abstract grade was also an ensemble model, achieving a mean average error (MAE) of 1.01.

**Conclusion:** While the baseline BOW model was the highest performing individual classifier, ensemble models that included state-of-the-art neural networks were able to outperform it. Our research shows that machine learning techniques can, to a reasonable degree of accuracy, predict both objective factors such as abstract category as well as subjective factors such as abstract grade. This work builds upon previous research involving using natural language processing on scientific abstracts to make useful inferences that address a meaningful problem.

## Introduction

With the advent of neural networks, automated analysis of arbitrary free text through natural language processing has become an area of active research in machine learning. In particular, recurrent and convolutional neural networks have been used on public data sets of biomedical literature to create algorithms for summarizing^1^, visualizing^2^, and analyzing large bodies of text. Additionally, there has been some work at attempting to predict the quality of body of work from the text alone. For example, Abrishami et al used deep neural network techniques to attempt to predict the number of citations on pieces of scientific literature using publicly available scientific literature as input^3^.

Currently, over 8000 abstracts are submitted to RSNA, the largest radiological society in North America, annually. For each abstract, human labor is required to assign or verify one of 16 top-level categories (e.g. “Breast Imaging”). Additionally, the reviewer assigns a numeric score from 1-10 based on a variety of subjective factors such as relevance, style and perceived writing quality. The process is manual, labor-intensive, and each abstract may receive input from multiple reviewers. The rationale for using neural networks for classifying and grading abstracts is twofold. First, if successful and reliable, an algorithm can save many hours of manual labor. Second, being able to assess literature quality and content has utility in research projects involving analyzing a large quantity of scientific literature.

Historically, a bag-of-words model was the standard for classifying free text^4^. Under this framework, only word frequencies are used as input to a machine learning model, thereby ignoring word order. Though naïve, the model has been shown to be difficult to outperform in previous works^5^. We hypothesize that with state-of-the-art neural network models involving techniques that consider word order, we can outperform a traditional bag-of-words model in both classifying and grading RSNA abstracts.

## Methods

A data set of 45527 abstracts was assembled using all RSNA abstract submissions from 2014 through 2019, inclusive, and associated metadata from manual review. Each abstract was tokenized using Stanford’s NLTK tokenizer^6^. Word endings were stripped using a stemmer from the same package. Preprocessing and vectorization were performed using Keras^7^, a Python library for deep learning. The data set was split into 60/20/20, with 60% of the data used for training algorithms, 20% used for real-time validation, and 20% used for final model testing and metrics reporting. The same split was used for all subsequent models.

A bag of words (BOW) model is a popular natural language processing model that discounts grammar and word ordering by only considering presence and frequency of words. One of many theoretical drawbacks of this model is the inability to detect negation. Nevertheless, based on the literature, BOW models perform quite well in practice and are difficult to outperform. As a baseline, a bag-of-words model was trained. First, a term-document matrix was generated by using a term frequency inverse document frequency (tf-idf) weighting scheme, a commonly used technique in text mining, natural language processing, and information retrieval^8^. With tf-idf, the importance of a word increases with the frequency that it is used, and decreases with the number of documents in which it can be found. Each abstract, with tf-idf weights, was represented into a single vector in the term-document matrix and placed into a single-layer-perceptron neural network model to evaluate a baseline for more sophisticated models. Weights and parameters were optimized using a grid search algorithm. Back-propagation with an Adam optimizer was used.

To test the hypothesis that more sophisticated models can surpass the performance of the baseline BOW model, individual words need to be vectorized. Each word in the corpus was embedded in a 250-dimensional space using GloVe, a validated software tool for generating word embeddings^9^. After attempting several other tools, GloVe was chosen for superior performance.

A convolutional neural network model was trained with sequential word embeddings as input^10^. The model is used widely in computer vision tasks, but has recently seen popularity in natural language processing, even outperforming other neural networks in certain classification tasks^11^. With a convolutional neural network, words adjacent to each other in the original text are pooled together before evaluation. Because of its ability to identify local context, word order and negation can be identified in certain cases, solving a theoretical drawback of the bag of words model. For the dataset, 1D convolutions with 128×5 kernel size and 1D max pooling was used. Parameters were optimized using a grid search algorithm. Metrics were then compared with the original bag of words model.

One drawback of a convolutional neural network is its inability to address relations between words across long distances because it would exceed the kernel size. Recurrent neural networks aim to solve this problem by taking word embedding inputs sequentially. The most common neural network cells used in these networks are long short-term memory cells (LSTM) ^12^ and gated recurrent units (GRU) ^13^. Both unit types within a neural network selectively “remember” relevant values over large spans of data. After evaluating both types of units, no significant differences were found. However, gated recurrent units had faster performance and were thus used for this project. A multi-layer recurrent neural network was generated with gated recurrent units, the model was compared to the bag of words model and the convolutional neural network model.

Finally, ensemble models have been shown to outperform single models in a wide array of domains^14^. In this paradigm, multiple models output a single “vote” towards a final result, with the final vote count used to determine the final classification. All three models (BOW, CNN, and GRU) were used in an ensemble model and metrics from that model were evaluated.

## Results

The baseline BOW model was able to achieve 71.8% overall accuracy on classifying categories and had a mean average error (MAE) of 1.06 on predicting the numerical grade given by the reviewer. Accuracy was different among different categories, with breast imaging having the highest accuracy at 93.7% and informatics the lowest at 20.5%.

Overall, both the CNN and GRU underperformed with respect to the BOW baseline model. The CNN achieved an overall accuracy of 64.4% on predicting categories, and MAE of 1.10 on abstract grade. The CNN achieved the highest accuracy on the “Informatics” category, and was the highest performing classifier overall in that category. The GRU model achieved an overall accuracy of 65.7% on predicting categories, and a MAE Of 1.29 on predicting grades. The GRU classifier performed the best on the “Pediatrics” category, and was the highest performing classifier overall in that category. On “Breast Imaging”, “Cardiac Imaging”, and “Musculoskeletal Imaging”, the baseline BOW model significantly outperformed more sophisticated models.

The ensemble model outperformed all other models and achieved both the highest accuracy in category classification and lowest MAE in predicting abstract grades. It achieved a 73.0% overall accuracy on classifying abstract category, also achieving the highest accuracy in the “Cardiac Imaging” category. In other categories, individual classifiers outperformed the ensemble model, though were overall less accurate. With respect to predicting abstract grades, the ensemble model had the lowest overall MAE as well as the lowest MAE in every individual category of abstract.

Table 1 shows a breakdown of the performance of BOW, CNN, GRU and ensemble models for classifying abstract category. Table 2 shows the breakdown of all models for classifying abstract grade.

**Table 1:** Accuracy of neural network classifiers on RSNA abstract category.

|  | BOW | CNN | GRU | Ensemble |
| --- | --- | --- | --- | --- |
| **Overall** | 71.8% | 64.4% | 65.7% | 73.0% |
| Breast | 93.7% | 76.1% | 88.5% | 93.0% |
| Cardiac | 86.4% | 69.3% | 80.0% | 87.1% |
| Informatics | 20.5% | 36.3% | 27.8% | 26.3% |
| Pediatrics | 48.2% | 51.5% | 57.7% | 56.1% |
| Physics | 53.6% | 55.9% | 43.1% | 54.4% |
| Musculoskeletal | 79.7% | 65.8% | 66.7% | 79.0% |

**Table 2:** Mean average error of neural network classifiers on RSNA abstract grade.

|  | BOW | CNN | GRU | Ensemble |
| --- | --- | --- | --- | --- |
| **Overall** | 1.06 | 1.10 | 1.29 | 1.01 |
| Breast | 1.00 | 1.09 | 1.20 | 0.94 |
| Cardiac | 0.91 | 0.94 | 1.12 | 0.85 |
| Informatics | 1.03 | 1.14 | 1.22 | 1.02 |
| Pediatrics | 1.12 | 1.24 | 1.32 | 1.09 |
| Physics | 1.03 | 1.08 | 1.27 | 1.01 |
| Musculoskeletal | 1.32 | 1.27 | 1.58 | 1.26 |

## Discussion

The single best classifier for abstract categories was the baseline BOW model. The literature supports that despite its simplicity and naïve approach to classifying texts, the BOW model is difficult in practice to outperform. One reason behind the superiority of the baseline model is that more complex models are prone to overfitting. When models are overly tuned to perform well in the training set, it may lose generalizability in the testing set. The BOW model did particularly well in categories such as “Breast Imaging”, where, upon manual inspection, a model could theoretically achieve a high accuracy simply by looking for the presence of certain high-yield words such as “Breast” or “Mammography”. More sophisticated models may attempt to look for more complex patterns that may be less specific for the category.

However, there were some cases where other models outperformed the BOW model. Notably, in the “Informatics” category, BOW did relatively poorly with an accuracy of only 20.%. By manually inspecting the documents, it appears that there is no universal set of words that would indicate that an abstract belongs to this category. Furthermore, many abstracts that contain the word “informatics” do not belong in this category, and simply use computation as part of an overall larger project. Of all the models, BOW had the lowest performance in this category. More sophisticated models can, in theory, detect multi-word features and/or features related to word ordering that allow for better classification results in this category.

Given the relative strengths and weaknesses of each model, it is intuitive that an ensemble model would outperform the baseline BOW model. Based on the literature, ensemble models tend to perform well when the underlying mechanism behind each model is distinct^15^. Even though the ensemble model was not the best classifier in every category, it performed significantly better than the baseline model overall.

With respect to abstract grades, all models performed well, with a MAE of around 1. The ensemble model outperformed all other models in this task, and unlike with abstract category classification, it outperformed all models in every category as well. Given the subjective nature of abstract grading and the relatively large number of factors involved in assigning a quality score, it is surprising that an algorithmic approach would do well. Both manual inspection and tree-based classifiers suggest that there are no particular words in an abstract that indicate whether it would receive a high or low score.

Previous work in the space has illustrated the ability of state-of-the-art neural networks to analyze scientific literature. Specific tasks include the ability to predict citation number from the text of the work^16^ or inferring aggregate information about specific entities mentioned in scientific literature^17^. However, no public work has been done in using neural networks to predict manually tagged categories and grades because public data sets of such information are not widely available. This work uses existing techniques on a closed-source data set and illustrates the feasibility of producing an algorithm that achieves reasonable accuracy in tagging and grading RSNA abstracts.

One limitation of this study is that while 73% accuracy is significantly higher than random guessing, it is not high enough to be used in a production environment. Further increases in accuracy through exploration of novel algorithms or further cleaning of the original dataset may be needed. Another limitation is that all models used are essentially “black boxes”. In making predictions, the neural network is unable to explain why it is making certain inferences. Using attention networks in the future can better elucidate underlying reasons why predictions are being made^18^.

## Data Availability

The data that support the findings of this study are available from RSNA. Restrictions apply to the availability of these data, which were used under license for this study.

## Acknowledgements

Thanks to Dr. Charles Kahn, of RSNA, for supplying the data set used for analysis in this work.

